# Cholesterol management gaps before a recurrent coronary event: insights from the Get With The Guidelines – Coronary Artery Disease registry

**DOI:** 10.64898/2026.03.19.26348857

**Authors:** Lisandro D. Colantonio, Zhixin Wang, Stephen L. Sigal, Emily B. Levitan, Vera Bittner

## Abstract

**Background:** Guidelines recommend that patients with coronary artery disease (CAD) lower their low-density lipoprotein cholesterol (LDL-C) using maximally tolerated statin therapy to prevent recurrent events.

**Methods:** We analyzed the prevalence of statin nonuse and an LDL-C ≥70 mg/dL in patients ≥18 years of age in the Get With The Guidelines-CAD registry with known CAD who were hospitalized for a new myocardial infarction or unstable angina in 2023-2024. Data collection on statin use and LDL-C at arrival is optional in the registry.

**Results:** Among 34,003 patients included (mean age 68 years; 71% male; 73% white), 31.6% did not use a statin before admission. The prevalence of statin nonuse was higher in women than in men (adjusted prevalence ratio [aPR] 1.08; 95% confidence interval [95%CI] 1.04, 1.14). LDL-C was not documented in 30.7% of patients. Among patients not using and using a statin, 74.6% and 49.8%, respectively, had an LDL-C ≥70 mg/dL. Women were more likely than men to have an LDL-C ≥70 mg/dL, whether using or not using a statin (aPR 1.18 [95%CI 1.13, 1.24] and 1.08 [95%CI 1.04, 1.12], respectively). Black and Hispanic patients were more likely to have an LDL-C ≥70 mg/dL compared to their white counterparts (aPR 1.30 [95%CI 1.24, 1.37] and 1.11 [95%CI 1.03, 1.19], respectively) among those using a statin. There were no statistically significant differences in LDL-C by race/ethnicity among those not using a statin.

**Conclusion:** Targeted quality improvement initiatives are needed to address ambulatory cholesterol treatment gaps in patients with known CAD.

**Clinical Perspective:** *What Is New?:*

- In a contemporary national registry of patients with known coronary artery disease hospitalized for recurrent acute coronary syndromes, about one-third were not using statin therapy prior to admission.
- Approximately three-quarters of statin nonusers and one-half of statin users had an LDL-C level ≥70 mg/dL at admission, indicating substantial residual risk despite current guideline recommendations.
- Women and Black and Hispanic patients were more likely to have inadequately controlled LDL-C, particularly among those already receiving statin therapy.

*What Are the Clinical Implications?:*

- Preventable ambulatory cholesterol treatment gaps frequently occur before recurrent coronary events, underscoring the need for more guideline-recommended outpatient lipid management.
- Routine LDL-C monitoring and timely intensification of lipid-lowering therapy, including high-intensity statins and add-on therapies when indicated, should be prioritized after coronary events.
- Targeted quality improvement strategies in the ambulatory setting are needed to address persistent cholesterol treatment gaps in secondary prevention care, including sex- and race/ethnicity-related disparities.

## Introduction

The 2018 American Heart Association (AHA)/American College of Cardiology (ACC)/multi-society cholesterol guideline and the 2025 ACC/AHA/multi-society guideline on acute coronary syndrome recommend maximally-tolerated statin therapy for patients with coronary artery disease (CAD) to lower their low-density lipoprotein cholesterol (LDL-C) and prevent recurrent events.^1,2^ Recurrent coronary events are associated with a high cost and healthcare services utilization, and a risk of death and disability.^3,4^ Therefore, CAD patients not using a statin or with high LDL-C who experience a recurrent event represent a missed opportunity to improve cardiovascular health in this population.

Over the past decades, there has been a substantial increase in the prescription of high-intensity statin therapy following discharge in patients with a CAD hospitalization.^5,6^ Despite this improvement, Aggarwal et al. reported that 32.1% of US adults with known CAD were not using a statin in 2015-2020, and that 73.5% had an LDL-C ≥70 mg/dL.^7^ However, these results may primarily reflect clinical practices before the release of the 2018 AHA/ACC/multi-society cholesterol guideline.

The purpose of the current study was to assess cholesterol treatment gaps (i.e., nonuse of statins and LDL-C ≥70 mg/dL) in patients with known CAD who were hospitalized for a recurrent acute coronary event using contemporary data from the Get With The Guidelines (GWTG) - CAD registry. GWTG, initiated by the AHA in 2000, is the largest quality improvement initiative for inpatient management of cardio and neurovascular disease in the US.^8,9^ Prior studies suggest that cholesterol management gaps differ by sex and race/ethnicity.^10–13^ Therefore, we investigated whether disparities in cholesterol treatment by sex and race/ethnicity persist after the publication of the 2018 AHA/ACC/multi-society cholesterol guideline.

## Methods

We conducted a retrospective analysis using data from the GWTG-CAD registry. The GWTG-CAD registry includes patients hospitalized with a ST-segment elevation myocardial infarction (STEMI), non-ST-segment elevation myocardial infarction (NSTEMI), unstable angina, or cardiac or non-cardiac chest pain at participating hospitals across all US census regions. Data in the GWTG-CAD registry are systematically collected by each participating hospital using an online, interactive registry tool powered by IQVIA (Cambridge, MA).

Data collection on the use of statins and other cholesterol-lowering medications prior to admission started in 2023. For the current study, we analyzed data from patients ≥18 years of age with known CAD who were hospitalized for a STEMI, NSTEMI, or unstable angina in 2023-2024. Known CAD was defined by a prior myocardial infarction, coronary artery bypass grafting (CABG), or percutaneous coronary intervention (PCI) in the patient’s medical history. We excluded patients whose symptoms were first identified in an acute care setting (including inpatient or emergency care at the same hospital or another) or in a chronic care facility, as these may be more likely to be receiving guideline-recommended therapy. We also excluded patients on dialysis, as lipid management in this population is guided by specific recommendations.^14^ Finally, we restricted the analysis to patients with data on cholesterol-lowering medication use at admission.

All institutions participating in the GWTG program are required to comply with privacy regulations, and all data enterers received appropriate training. The current analysis was approved by the Institutional Review Board of the University of Alabama at Birmingham. De-identified GWTG-CAD data were obtained from the AHA through the Precision Medicine Platform (https://pmp.heart.org/).^15^

### Study variables

The outcome variables for the current analysis include the nonuse of statins prior to admission and LDL-C at arrival. Nonuse of statins, ezetimibe, or proprotein convertase subtilisin/kexin type 9 inhibitors (PCSK9i) was examined as a secondary outcome. Data collection on prior cholesterol-lowering medication use and LDL-C at arrival is optional in the GWTG-CAD registry. Information on the intensity of prior statin therapy is not available.

Socio-demographic variables analyzed in the current study include year of hospitalization, age at admission, sex at birth, race/ethnicity (patient-defined and analyzed as a social construct), payment source, AHA geographic region, and health-related unmet social needs. Government insurance was defined as having at least one of the following payment sources: Medicare, Medicaid, Veterans Affairs, Civilian Health and Medical Program of the Department of Veterans Affairs (CHAMPVA), Tricare, and Indian Health Services. Private insurance includes commercial insurance, typically tied to employer-based plans. Having unmet social needs was defined by reported gaps in any of the following health-related areas: education, employment, financial strain, food, living situation/housing, mental health, personal safety, substance use disorder, transportation barriers, and utilities.

Clinical variables analyzed include body mass index (calculated using weight and height at arrival), history of smoking in the past 12 months, and medical history (i.e., history of stroke, peripheral artery disease [PAD], heart failure, cancer, prior CABG, prior PCI, diabetes, dyslipidemia, and hypertension). Discharge variables analyzed include disposition and prescription of any statin and, separately, high-intensity statin therapy.

Patient-level data in the GWTG-CAD registry have been linked to hospital data. Hospital-level data used in the current study include bed size, accreditation by The Joint Commission, heart-related services (i.e., adult cardiac surgery, adult cardiology, adult diagnostic catheterization, and adult interventional cardiac catheterization), and rural referral center, sole community provider, and teaching status. Major teaching hospitals are defined as those with a Council of Teaching Hospitals designation. Minor teaching hospitals are those approved to participate in residency and/or internship training by the Accreditation Council for Graduate Medical Education or the American Osteopathic Association, and those with medical school affiliation reported to the American Medical Association. All other hospitals are classified as non-teaching.

### Statistical analysis

We calculated descriptive statistics for characteristics of patients using and not using a statin at admission. We also calculated the proportion of patients not using a statin at admission, overall and by sex and race/ethnicity, separately. We used generalized estimating equation Poisson regression with robust variance estimators to obtain prevalence ratios (PR) for statin nonuse at admission by sex and race/ethnicity, accounting for clustering within hospitals.^16^ Model 1 was unadjusted. Model 2 included sex and race/ethnicity, adjusted for calendar year, age, and AHA geographic region. A fully adjusted model, Model 3, included sex and race/ethnicity, adjusted for all socio-demographic, clinical, and hospital-level variables. Model 3 also included interaction terms between sex and racial/ethnic groups to determine whether sex disparities differ by race/ethnicity. Few patients take ezetimibe or a PCSK9i.^7,17^ In a sensitivity analysis, we repeated the procedures above to analyze the proportion of patients not using a statin, ezetimibe, or a PCSK9i at admission. Finally, among patients who were discharged alive to their homes, we calculated the proportion who were not prescribed any statin and, separately, high-intensity statin therapy, overall and by sex and race/ethnicity. We used generalized estimating equation Poisson regression as described above to compare the prescription of statin therapy at discharge by sex and race/ethnicity.

We calculated descriptive statistics for patients with an LDL-C ≥70 mg/dL and <70 mg/dL. We also calculated the proportion of patients with an LDL-C ≥70 mg/dL, overall, and by statin use at admission. We compared the proportion of patients with an LDL-C ≥70 mg/dL by sex and race/ethnicity, separately, stratified by prior statin use. We used regression Models 1-3 described above to obtain PRs for having an LDL-C ≥70 mg/dL associated with statin use, sex, and race/ethnicity.^16^ There are sex and racial/ethnic differences in the use of ezetimibe and PCSK9i, which may be a confounder for LDL-C levels.^17^ Therefore, Model 3 included further adjustment for ezetimibe and PCSK9i use. In sensitivity analyses, we analyzed the proportion of patients with an LDL-C ≥100 mg/dL, based on the last performance measure for cholesterol management in patients with CAD included in the AHA/ACC Report on Performance Measures and the Healthcare Effectiveness Data and Information Set (HEDIS), which was retired in 2015.^18–20^ We also analyzed the proportion of patients with an LDL-C ≥55 mg/dL, as an LDL-C goal of <55 mg/dL is recommended by European cholesterol management guidelines.^21,22^

In exploratory analyses, we identified socio-demographic, clinical, and hospital-level characteristics associated with statin nonuse and, separately, an LDL-C ≥70 mg/dL at arrival. All analyses were conducted in the Precision Medicine Platform using R Studio and SAS (SAS Institute Inc., Cary NC).^15^

## Results

The analysis included 34,003 patients from 666 hospitals (**Figure 1**), of whom 10,738 (31.6%) did not use a statin before admission. Patients not using a statin were younger and more likely to be from the Midwest, Western, Southeast, and Southwest versus the Eastern region, and to be uninsured compared to those using a statin (**Table 1**). Patients not using a statin were less likely to have a history of PAD, heart failure, CABG, PCI, diabetes, and dyslipidemia.

**Figure 1.**
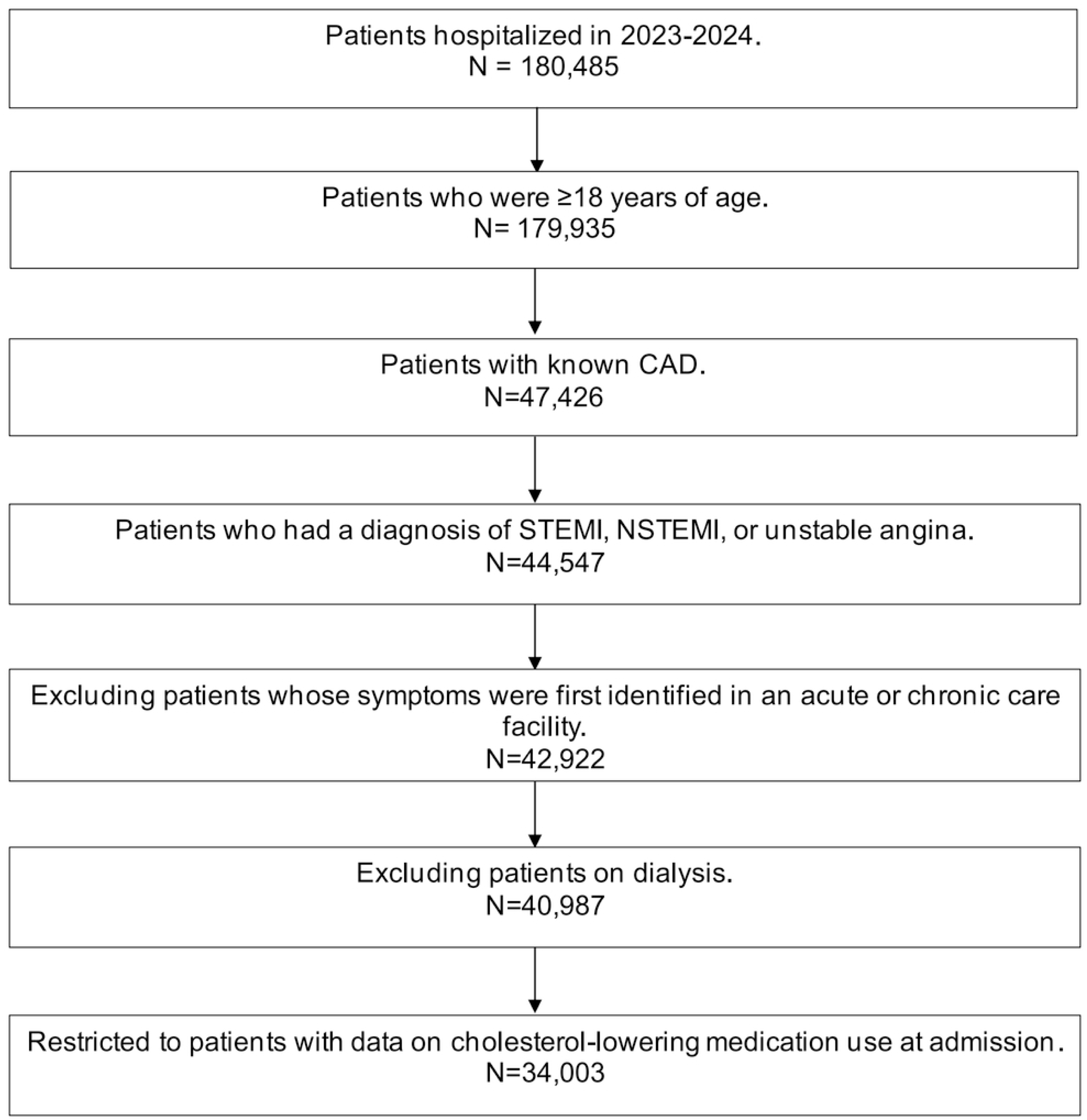
Flow-chart of GWTG-CAD patients included in the current analysis. CAD: coronary artery disease; GWTG: Get With The Guidelines; NSTEMI: non-ST-segment elevation myocardial infarction; STEMI: ST-segment elevation myocardial infarction.

**Table 1.**
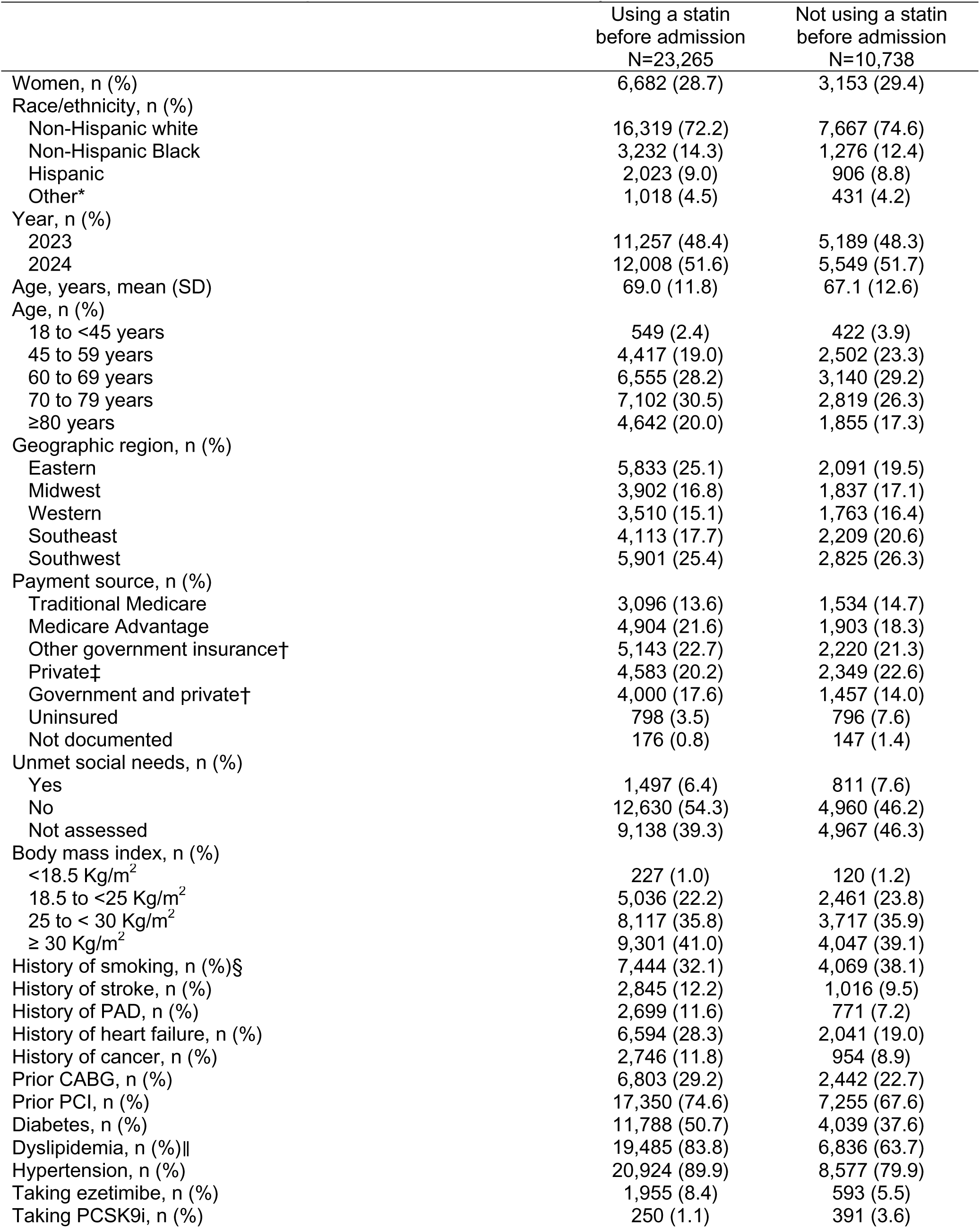
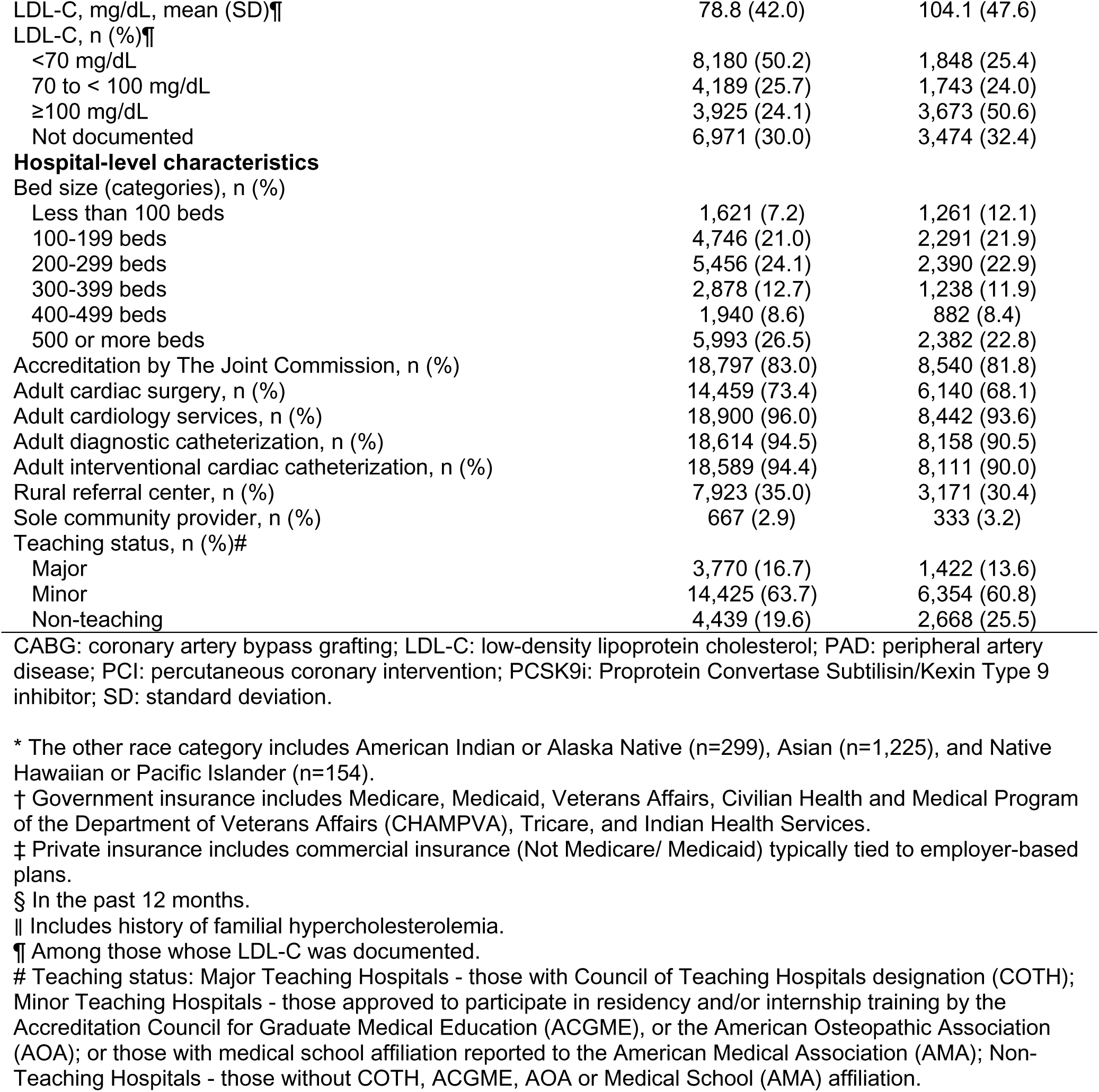
Characteristics of patients included in the analysis.

|  | Using a statin<br>before admission<br>N=23,265 | Not using a statin<br>before admission<br>N=10,738 |
| --- | --- | --- |
| Women, n (%) | 6,682 (28.7) | 3,153 (29.4) |
| Race/ethnicity, n (%) |  |  |
| Non-Hispanic white | 16,319 (72.2) | 7,667 (74.6) |
| Non-Hispanic Black | 3,232 (14.3) | 1,276 (12.4) |
| Hispanic | 2,023 (9.0) | 906 (8.8) |
| Other* | 1,018 (4.5) | 431 (4.2) |
| Year, n (%) |  |  |
| 2023 | 11,257 (48.4) | 5,189 (48.3) |
| 2024 | 12,008 (51.6) | 5,549 (51.7) |
| Age, years, mean (SD) | 69.0 (11.8) | 67.1 (12.6) |
| Age, n (%) |  |  |
| 18 to <45 years | 549 (2.4) | 422 (3.9) |
| 45 to 59 years | 4,417 (19.0) | 2,502 (23.3) |
| 60 to 69 years | 6,555 (28.2) | 3,140 (29.2) |
| 70 to 79 years | 7,102 (30.5) | 2,819 (26.3) |
| ≥80 years | 4,642 (20.0) | 1,855 (17.3) |
| Geographic region, n (%) |  |  |
| Eastern | 5,833 (25.1) | 2,091 (19.5) |
| Midwest | 3,902 (16.8) | 1,837 (17.1) |
| Western | 3,510 (15.1) | 1,763 (16.4) |
| Southeast | 4,113 (17.7) | 2,209 (20.6) |
| Southwest | 5,901 (25.4) | 2,825 (26.3) |
| Payment source, n (%) |  |  |
| Traditional Medicare | 3,096 (13.6) | 1,534 (14.7) |
| Medicare Advantage | 4,904 (21.6) | 1,903 (18.3) |
| Other government insurance† | 5,143 (22.7) | 2,220 (21.3) |
| Private‡ | 4,583 (20.2) | 2,349 (22.6) |
| Government and private† | 4,000 (17.6) | 1,457 (14.0) |
| Uninsured | 798 (3.5) | 796 (7.6) |
| Not documented | 176 (0.8) | 147 (1.4) |
| Unmet social needs, n (%) |  |  |
| Yes | 1,497 (6.4) | 811 (7.6) |
| No | 12,630 (54.3) | 4,960 (46.2) |
| Not assessed | 9,138 (39.3) | 4,967 (46.3) |
| Body mass index, n (%) |  |  |
| <18.5 Kg/m<sup>2</sup> | 227 (1.0) | 120 (1.2) |
| 18.5 to <25 Kg/m<sup>2</sup> | 5,036 (22.2) | 2,461 (23.8) |
| 25 to < 30 Kg/m<sup>2</sup> | 8,117 (35.8) | 3,717 (35.9) |
| ≥ 30 Kg/m<sup>2</sup> | 9,301 (41.0) | 4,047 (39.1) |
| History of smoking, n (%)§ | 7,444 (32.1) | 4,069 (38.1) |
| History of stroke, n (%) | 2,845 (12.2) | 1,016 (9.5) |
| History of PAD, n (%) | 2,699 (11.6) | 771 (7.2) |
| History of heart failure, n (%) | 6,594 (28.3) | 2,041 (19.0) |
| History of cancer, n (%) | 2,746 (11.8) | 954 (8.9) |
| Prior CABG, n (%) | 6,803 (29.2) | 2,442 (22.7) |
| Prior PCI, n (%) | 17,350 (74.6) | 7,255 (67.6) |
| Diabetes, n (%) | 11,788 (50.7) | 4,039 (37.6) |
| Dyslipidemia, n (%)\|\| | 19,485 (83.8) | 6,836 (63.7) |
| Hypertension, n (%) | 20,924 (89.9) | 8,577 (79.9) |
| Taking ezetimibe, n (%) | 1,955 (8.4) | 593 (5.5) |
| Taking PCSK9i, n (%) | 250 (1.1) | 391 (3.6) |
| Colantonio – Cholesterol treatment gaps GWTG |  |  |
| LDL-C, mg/dL, mean (SD)¶ | 78.8 (42.0) | 104.1 (47.6) |
| LDL-C, n (%)¶ |  |  |
| <70 mg/dL | 8,180 (50.2) | 1,848 (25.4) |
| 70 to < 100 mg/dL | 4,189 (25.7) | 1,743 (24.0) |
| ≥100 mg/dL | 3,925 (24.1) | 3,673 (50.6) |
| Not documented | 6,971 (30.0) | 3,474 (32.4) |
| **Hospital-level characteristics** |  |  |
| Bed size (categories), n (%) |  |  |
| Less than 100 beds | 1,621 (7.2) | 1,261 (12.1) |
| 100-199 beds | 4,746 (21.0) | 2,291 (21.9) |
| 200-299 beds | 5,456 (24.1) | 2,390 (22.9) |
| 300-399 beds | 2,878 (12.7) | 1,238 (11.9) |
| 400-499 beds | 1,940 (8.6) | 882 (8.4) |
| 500 or more beds | 5,993 (26.5) | 2,382 (22.8) |
| Accreditation by The Joint Commission, n (%) | 18,797 (83.0) | 8,540 (81.8) |
| Adult cardiac surgery, n (%) | 14,459 (73.4) | 6,140 (68.1) |
| Adult cardiology services, n (%) | 18,900 (96.0) | 8,442 (93.6) |
| Adult diagnostic catheterization, n (%) | 18,614 (94.5) | 8,158 (90.5) |
| Adult interventional cardiac catheterization, n (%) | 18,589 (94.4) | 8,111 (90.0) |
| Rural referral center, n (%) | 7,923 (35.0) | 3,171 (30.4) |
| Sole community provider, n (%) | 667 (2.9) | 333 (3.2) |
| Teaching status, n (%)# |  |  |
| Major | 3,770 (16.7) | 1,422 (13.6) |
| Minor | 14,425 (63.7) | 6,354 (60.8) |
| Non-teaching | 4,439 (19.6) | 2,668 (25.5) |

CABG: coronary artery bypass grafting; LDL-C: low-density lipoprotein cholesterol; PAD: peripheral artery disease; PCI: percutaneous coronary intervention; PCSK9i: Proprotein Convertase Subtilisin/Kexin Type 9 inhibitor; SD: standard deviation.
\* The other race category includes American Indian or Alaska Native (n=299), Asian (n=1,225), and Native Hawaiian or Pacific Islander (n=154).
† Government insurance includes Medicare, Medicaid, Veterans Affairs, Civilian Health and Medical Program of the Department of Veterans Affairs (CHAMPVA), Tricare, and Indian Health Services.
‡ Private insurance includes commercial insurance (Not Medicare/ Medicaid) typically tied to employer-based plans.
§ In the past 12 months.
|| Includes history of familial hypercholesterolemia.
¶ Among those whose LDL-C was documented.

Statin nonuse by sex and race/ethnicity is shown in **Table 2**. After full multivariable adjustment (Model 3), the PR for statin nonuse comparing women versus men was 1.08 (95% confidence interval [CI] 1.04, 1.14). The fully adjusted PR for statin nonuse comparing non-Hispanic Black versus non-Hispanic white patients was 0.91 (95% CI 0.83, 0.99). No other racial/ethnic differences were statistically significant after full adjustment. There was no evidence of sex-race/ethnicity interaction (p-value 0.30).

**Table 2.** Prevalence ratios for statin nonuse before admission associated with sex and race/ethnicity (n=34,003).

|  | Statin nonuse, n (%) | Prevalence ratio (95% CI) |  |  |
| --- | --- | --- | --- | --- |
|  |  | Model 1 | Model 2 | Model 3* |
| Overall | 10,738 (31.6) | --- | --- | --- |
| **Sex** |  |  |  |  |
| Men | 7,583 (31.4) | 1 (ref) | 1 (ref) | 1 (ref) |
| Women | 3,153 (32.1) | 1.02 (0.99, 1.06) | 1.06 (1.02, 1.10) | 1.08 (1.04, 1.14) |
| **Race/ethnicity** |  |  |  |  |
| Non-Hispanic white | 7,667 (32.0) | 1 (ref) | 1 (ref) | 1 (ref) |
| Non-Hispanic Black | 1,276 (28.3) | 0.89 (0.82, 0.95) | 0.84 (0.79, 0.91) | 0.91 (0.83, 0.99) |
| Hispanic | 906 (30.9) | 0.97 (0.89, 1.05) | 0.92 (0.85, 1.00) | 1.02 (0.92, 1.12) |
| Other | 431 (29.7) | 0.93 (0.84, 1.03) | 0.90 (0.82, 0.99) | 0.99 (0.89, 1.10) |

CABG: coronary artery bypass grafting; CI: confidence interval; PAD: peripheral artery disease; PCI: percutaneous coronary intervention.
Model 1 is crude.
Model 2 includes sex and race/ethnicity, simultaneously, with adjustment for calendar year, age, and geographic region.
Model 3 includes sex and race/ethnicity, with adjustment for calendar year, age, geographic region, payment source, unmet social needs, body mass index, history of smoking, stroke, PAD, heart failure, and cancer, prior CABG and PCI, diabetes, dyslipidemia, hypertension, and hospital-level characteristics, including bed size, accreditation by The Joint Commission, adult cardiac surgery, adult cardiology services, adult diagnostic catheterization, adult interventional cardiac catheterization, rural referral center, sole community provider, and teaching Status. Model 3 also includes interaction terms between sex and racial/ethnic groups to determine whether sex disparities differ by race/ethnicity.
\* p-value for the interaction between sex and racial/ethnic groups: 0.30.

In the sensitivity analysis evaluating prior use of a statin, ezetimibe, or PCSK9i, 9,836 patients (28.9%) were not taking any of these medications. There were no statistically significant differences in the use of a statin, ezetimibe, or PCSK9i by sex or race/ethnicity (**Supplemental Table 1**).

Among the 25,062 patients discharged alive to their homes, 6.4% were not prescribed a statin, and 14.7% were not prescribed high-intensity statin therapy. Nonprescription of a statin and high-intensity statin therapy were each more common in women than in men, and less common among non-Hispanic Black and Hispanic patients than their non-Hispanic white counterparts (**Supplemental Table 2**).

### LDL-C levels

Among 23,558 patients with LDL-C documented at arrival, 13,530 (57.4%) had an LDL-C ≥70 mg/dL. Patients with an LDL-C ≥70 mg/dL were younger and more likely to be women, non-Hispanic Black, and uninsured than those with an LDL-C <70 mg/dL (**Table 3**). Patients with an LDL-C ≥70 mg/dL were also less likely to have a prior CABG or diabetes. Patients without LDL-C documented were more likely to have been hospitalized in small (<100 beds), non-teaching hospitals than those with LDL-C documented (**Supplemental Table 3**).

**Table 3.**
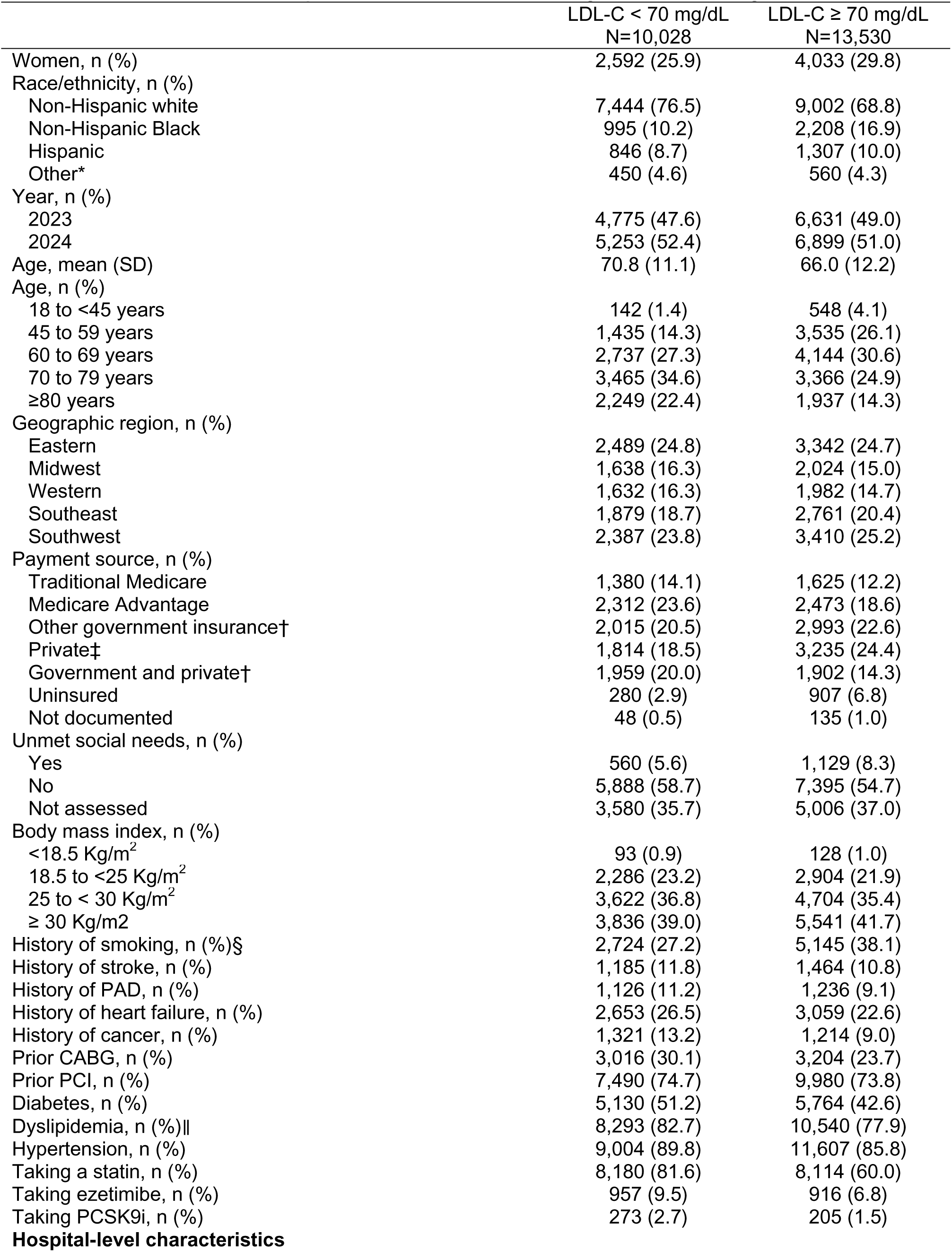
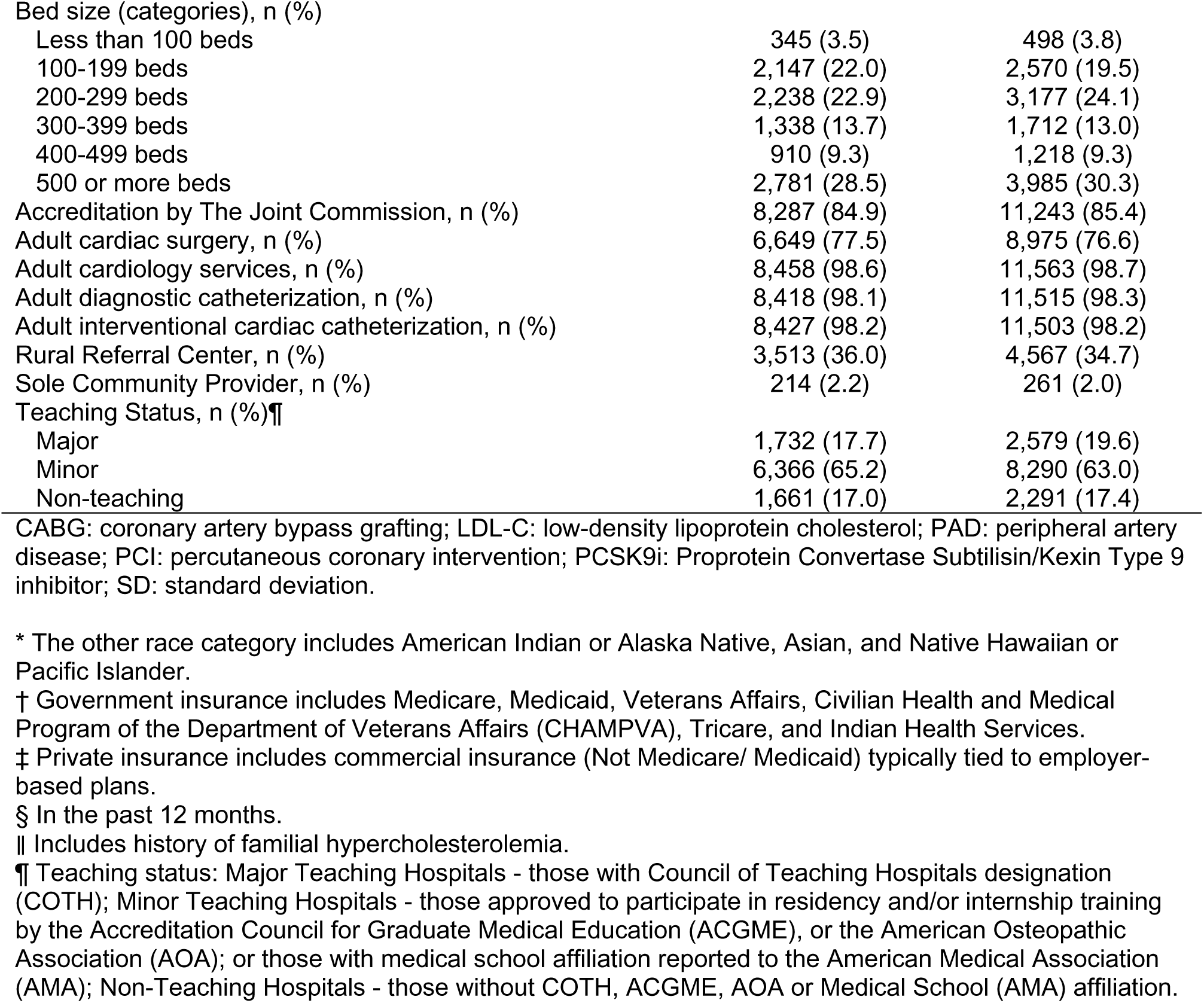
Characteristics of patients with LDL-C <70 mg/dL and ≥70 mg/dL.

|  | LDL-C < 70 mg/dL<br>N=10,028 | LDL-C ≥ 70 mg/dL<br>N=13,530 |
| --- | --- | --- |
| Women, n (%) | 2,592 (25.9) | 4,033 (29.8) |
| Race/ethnicity, n (%) |  |  |
| Non-Hispanic white | 7,444 (76.5) | 9,002 (68.8) |
| Non-Hispanic Black | 995 (10.2) | 2,208 (16.9) |
| Hispanic | 846 (8.7) | 1,307 (10.0) |
| Other* | 450 (4.6) | 560 (4.3) |
| Year, n (%) |  |  |
| 2023 | 4,775 (47.6) | 6,631 (49.0) |
| 2024 | 5,253 (52.4) | 6,899 (51.0) |
| Age, mean (SD) | 70.8 (11.1) | 66.0 (12.2) |
| Age, n (%) |  |  |
| 18 to <45 years | 142 (1.4) | 548 (4.1) |
| 45 to 59 years | 1,435 (14.3) | 3,535 (26.1) |
| 60 to 69 years | 2,737 (27.3) | 4,144 (30.6) |
| 70 to 79 years | 3,465 (34.6) | 3,366 (24.9) |
| ≥80 years | 2,249 (22.4) | 1,937 (14.3) |
| Geographic region, n (%) |  |  |
| Eastern | 2,489 (24.8) | 3,342 (24.7) |
| Midwest | 1,638 (16.3) | 2,024 (15.0) |
| Western | 1,632 (16.3) | 1,982 (14.7) |
| Southeast | 1,879 (18.7) | 2,761 (20.4) |
| Southwest | 2,387 (23.8) | 3,410 (25.2) |
| Payment source, n (%) |  |  |
| Traditional Medicare | 1,380 (14.1) | 1,625 (12.2) |
| Medicare Advantage | 2,312 (23.6) | 2,473 (18.6) |
| Other government insurance† | 2,015 (20.5) | 2,993 (22.6) |
| Private‡ | 1,814 (18.5) | 3,235 (24.4) |
| Government and private† | 1,959 (20.0) | 1,902 (14.3) |
| Uninsured | 280 (2.9) | 907 (6.8) |
| Not documented | 48 (0.5) | 135 (1.0) |
| Unmet social needs, n (%) |  |  |
| Yes | 560 (5.6) | 1,129 (8.3) |
| No | 5,888 (58.7) | 7,395 (54.7) |
| Not assessed | 3,580 (35.7) | 5,006 (37.0) |
| Body mass index, n (%) |  |  |
| <18.5 Kg/m<sup>2</sup> | 93 (0.9) | 128 (1.0) |
| 18.5 to <25 Kg/m<sup>2</sup> | 2,286 (23.2) | 2,904 (21.9) |
| 25 to < 30 Kg/m<sup>2</sup> | 3,622 (36.8) | 4,704 (35.4) |
| ≥ 30 Kg/m<sup>2</sup> | 3,836 (39.0) | 5,541 (41.7) |
| History of smoking, n (%)§ | 2,724 (27.2) | 5,145 (38.1) |
| History of stroke, n (%) | 1,185 (11.8) | 1,464 (10.8) |
| History of PAD, n (%) | 1,126 (11.2) | 1,236 (9.1) |
| History of heart failure, n (%) | 2,653 (26.5) | 3,059 (22.6) |
| History of cancer, n (%) | 1,321 (13.2) | 1,214 (9.0) |
| Prior CABG, n (%) | 3,016 (30.1) | 3,204 (23.7) |
| Prior PCI, n (%) | 7,490 (74.7) | 9,980 (73.8) |
| Diabetes, n (%) | 5,130 (51.2) | 5,764 (42.6) |
| Dyslipidemia, n (%)\|\| | 8,293 (82.7) | 10,540 (77.9) |
| Hypertension, n (%) | 9,004 (89.8) | 11,607 (85.8) |
| Taking a statin, n (%) | 8,180 (81.6) | 8,114 (60.0) |
| Taking ezetimibe, n (%) | 957 (9.5) | 916 (6.8) |
| Taking PCSK9i, n (%) | 273 (2.7) | 205 (1.5) |
| Colantonio – Cholesterol treatment gaps GWTG |  |  |
| Bed size (categories), n (%) |  |  |
| Less than 100 beds | 345 (3.5) | 498 (3.8) |
| 100-199 beds | 2,147 (22.0) | 2,570 (19.5) |
| 200-299 beds | 2,238 (22.9) | 3,177 (24.1) |
| 300-399 beds | 1,338 (13.7) | 1,712 (13.0) |
| 400-499 beds | 910 (9.3) | 1,218 (9.3) |
| 500 or more beds | 2,781 (28.5) | 3,985 (30.3) |
| Accreditation by The Joint Commission, n (%) | 8,287 (84.9) | 11,243 (85.4) |
| Adult cardiac surgery, n (%) | 6,649 (77.5) | 8,975 (76.6) |
| Adult cardiology services, n (%) | 8,458 (98.6) | 11,563 (98.7) |
| Adult diagnostic catheterization, n (%) | 8,418 (98.1) | 11,515 (98.3) |
| Adult interventional cardiac catheterization, n (%) | 8,427 (98.2) | 11,503 (98.2) |
| Rural Referral Center, n (%) | 3,513 (36.0) | 4,567 (34.7) |
| Sole Community Provider, n (%) | 214 (2.2) | 261 (2.0) |
| Teaching Status, n (%)¶ |  |  |
| Major | 1,732 (17.7) | 2,579 (19.6) |
| Minor | 6,366 (65.2) | 8,290 (63.0) |
| Non-teaching | 1,661 (17.0) | 2,291 (17.4) |

CABG: coronary artery bypass grafting; LDL-C: low-density lipoprotein cholesterol; PAD: peripheral artery disease; PCI: percutaneous coronary intervention; PCSK9i: Proprotein Convertase Subtilisin/Kexin Type 9 inhibitor; SD: standard deviation.
\* The other race category includes American Indian or Alaska Native, Asian, and Native Hawaiian or Pacific Islander.
† Government insurance includes Medicare, Medicaid, Veterans Affairs, Civilian Health and Medical Program of the Department of Veterans Affairs (CHAMPVA), Tricare, and Indian Health Services.
‡ Private insurance includes commercial insurance (Not Medicare/ Medicaid) typically tied to employer-based plans.
§ In the past 12 months.
|| Includes history of familial hypercholesterolemia.
¶ Teaching status: Major Teaching Hospitals - those with Council of Teaching Hospitals designation (COTH); Minor Teaching Hospitals - those approved to participate in residency and/or internship training by the Accreditation Council for Graduate Medical Education (ACGME), or the American Osteopathic Association (AOA); or those with medical school affiliation reported to the American Medical Association (AMA); Non-Teaching Hospitals - those without COTH, ACGME, AOA or Medical School (AMA) affiliation.

Among patients not using and using a statin, 74.6% and 49.8%, respectively, had an LDL-C ≥70 mg/dL (**Table 4**). After full multivariable adjustment, the PR for an LDL-C ≥70 mg/dL associated with statin nonuse was 1.46 (95% CI 1.42, 1.51). Women were more likely than men to have an LDL-C ≥70 mg/dL among both those using and not using a statin. Non-Hispanic Black and Hispanic patients were more likely to have an LDL-C ≥70 mg/dL than their non-Hispanic white counterparts among those using a statin. There were no statistically significant differences by race/ethnicity among statin nonusers.

**Table 4.**
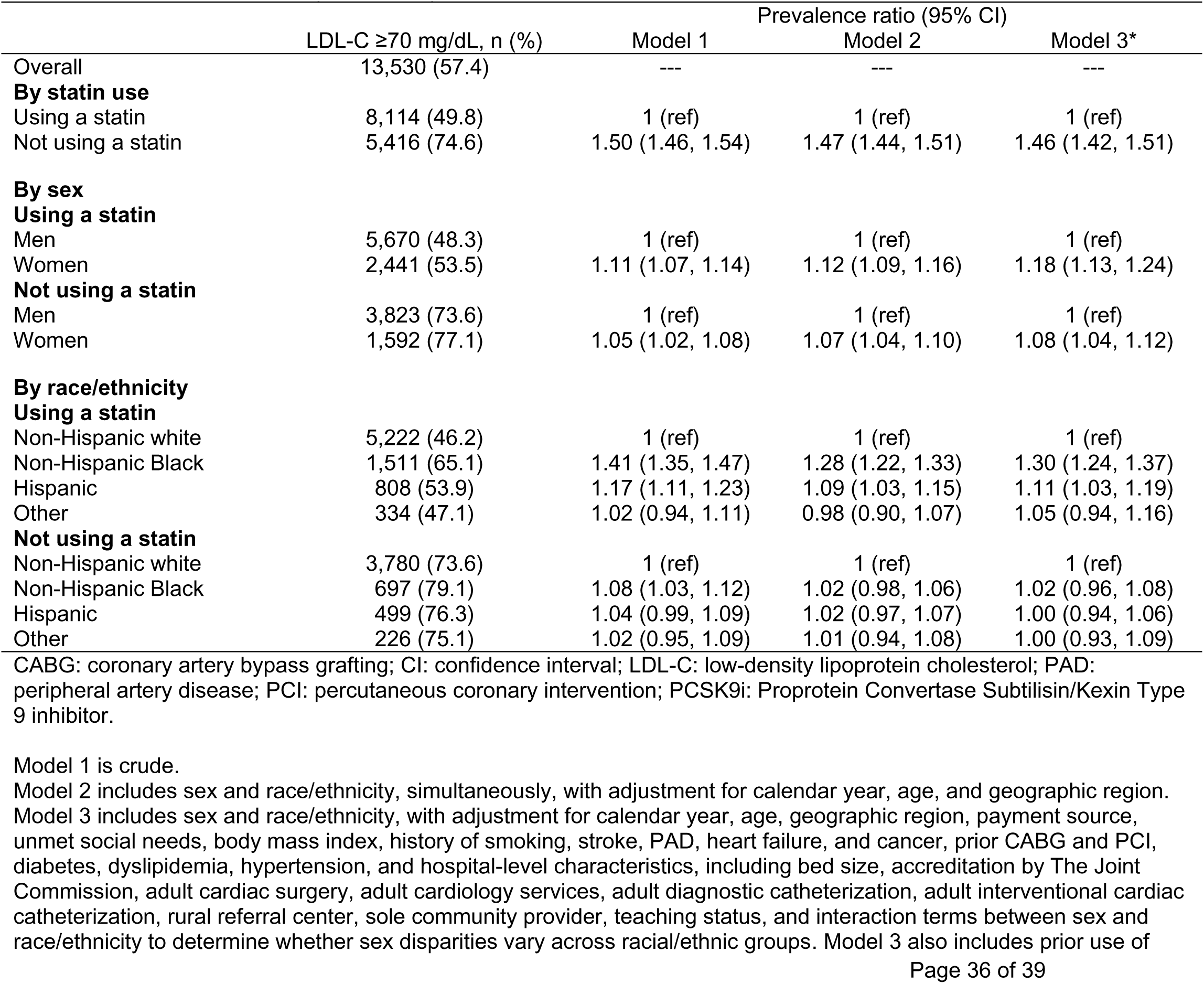
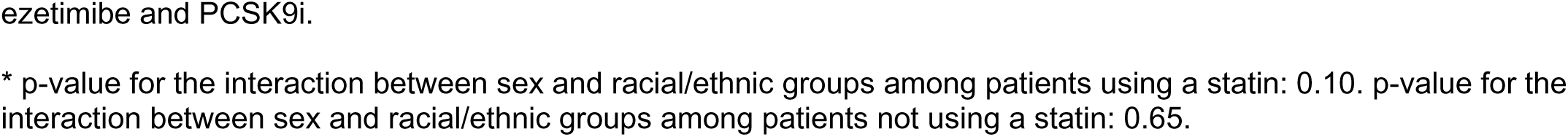
Prevalence ratios for an LDL-C ≥70 mg/dL associated with statin nonuse, sex, and race/ethnicity among patients with LDL-C documented (n=23,558).

In the sensitivity analysis, 32.3% of patients had an LDL-C ≥100 mg/dL, including 50.6% and 24.1% among those not using and using a statin, respectively (**Supplemental Table 4**). Sex disparities were larger than those observed in the main analysis (i.e., fully adjusted PR for an LDL-C ≥100 mg/dL associated with women among those using and not using a statin 1.32 [95% CI 1.22, 1.43] and 1.21 [95% CI 1.14, 1.29], respectively).

After full multivariable adjustment, the PR for an LDL-C ≥100 mg/dL comparing non-Hispanic Black versus non-Hispanic white patients using a statin was 1.37 (95% CI 1.25, 1.50). No other racial/ethnic differences were statistically significant after full adjustment among patients using and not using a statin. Overall, 74.6% of patients had an LDL-C ≥55 mg/dL (**Supplemental Table 5**).

### Exploratory analyses

After full multivariable adjustment, the PR for statin nonuse associated with being uninsured was 1.22 (95% CI 1.11, 1.34). Fully adjusted PRs associated with a history of heart failure, diabetes, and dyslipidemia were 0.79 (95% CI 0.75, 0.83), 0.80 (95% CI 0.77, 0.83), and 0.61 (95% CI 0.58, 0.64), respectively. All other PRs for statin nonuse ranged between 0.80 and 1.20 (**Table 5**).

**Table 5.**
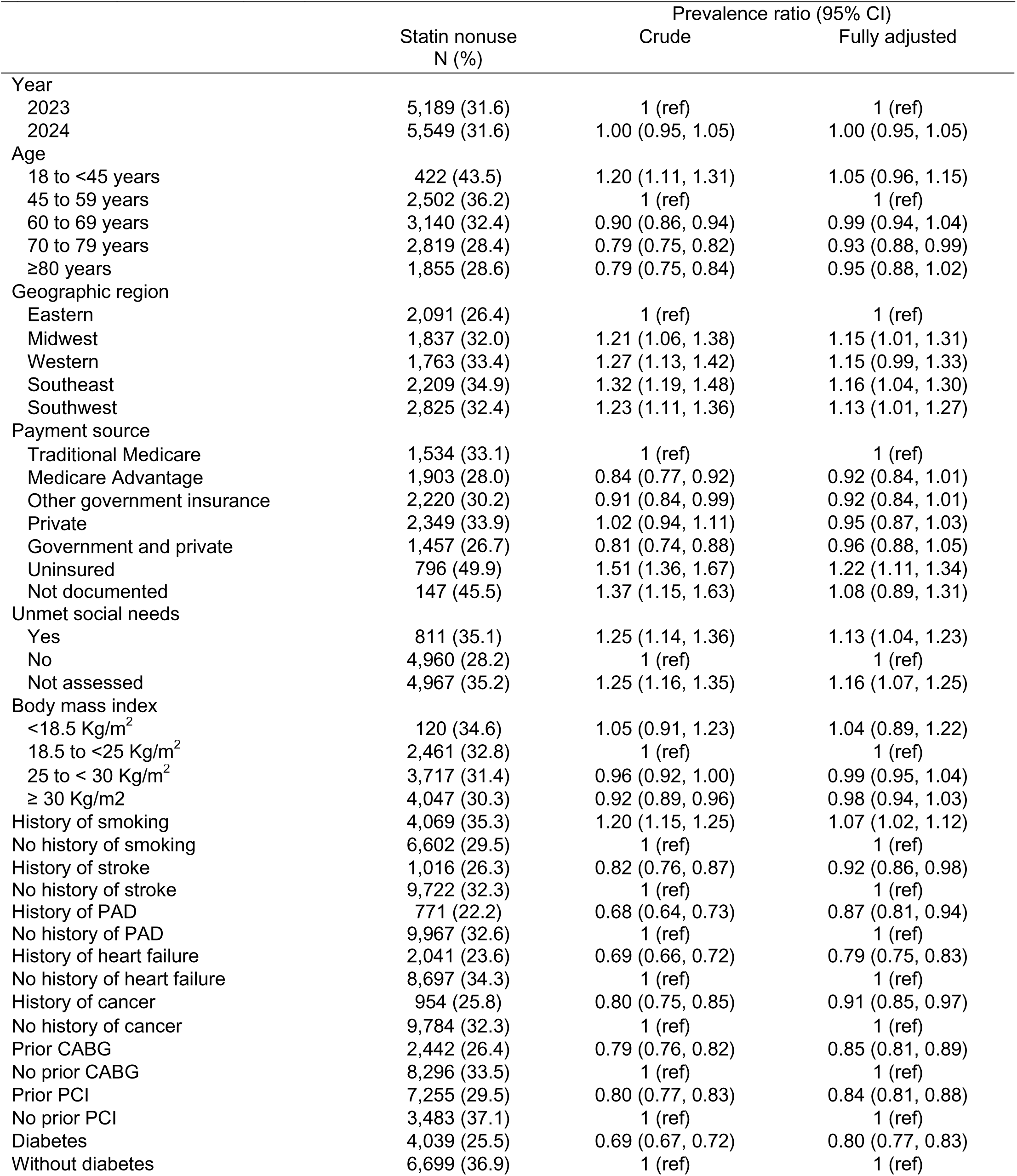
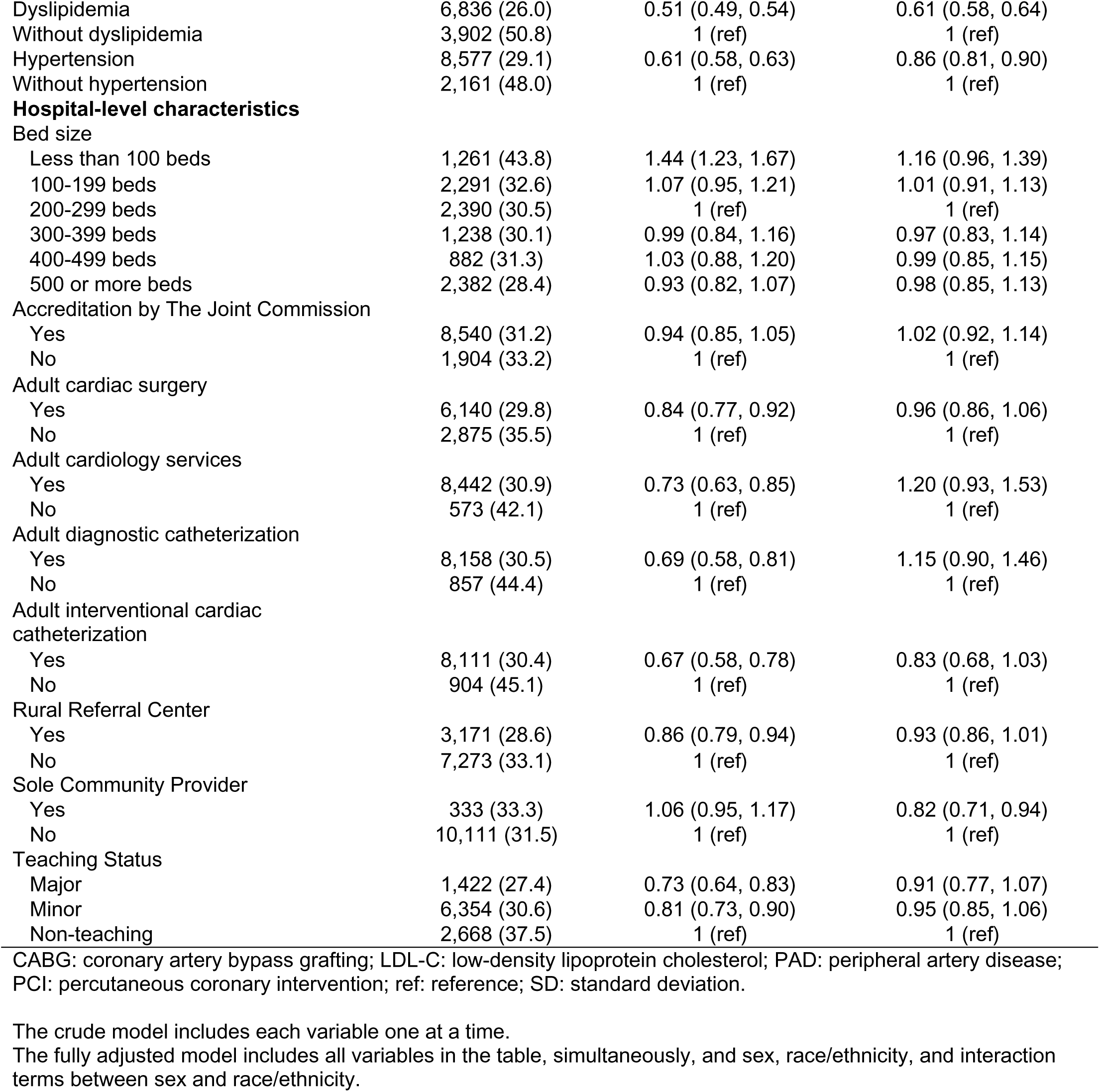
Patient characteristics associated with statin nonuse before admission (n=34,003). Exploratory analysis.

|  | Statin nonuse<br>N (%) | Prevalence ratio (95% CI) |  |
| --- | --- | --- | --- |
|  |  | Crude | Fully adjusted |
| Year |  |  |  |
| 2023 | 5,189 (31.6) | 1 (ref) | 1 (ref) |
| 2024 | 5,549 (31.6) | 1.00 (0.95, 1.05) | 1.00 (0.95, 1.05) |
| Age |  |  |  |
| 18 to <45 years | 422 (43.5) | 1.20 (1.11, 1.31) | 1.05 (0.96, 1.15) |
| 45 to 59 years | 2,502 (36.2) | 1 (ref) | 1 (ref) |
| 60 to 69 years | 3,140 (32.4) | 0.90 (0.86, 0.94) | 0.99 (0.94, 1.04) |
| 70 to 79 years | 2,819 (28.4) | 0.79 (0.75, 0.82) | 0.93 (0.88, 0.99) |
| ≥80 years | 1,855 (28.6) | 0.79 (0.75, 0.84) | 0.95 (0.88, 1.02) |
| Geographic region |  |  |  |
| Eastern | 2,091 (26.4) | 1 (ref) | 1 (ref) |
| Midwest | 1,837 (32.0) | 1.21 (1.06, 1.38) | 1.15 (1.01, 1.31) |
| Western | 1,763 (33.4) | 1.27 (1.13, 1.42) | 1.15 (0.99, 1.33) |
| Southeast | 2,209 (34.9) | 1.32 (1.19, 1.48) | 1.16 (1.04, 1.30) |
| Southwest | 2,825 (32.4) | 1.23 (1.11, 1.36) | 1.13 (1.01, 1.27) |
| Payment source |  |  |  |
| Traditional Medicare | 1,534 (33.1) | 1 (ref) | 1 (ref) |
| Medicare Advantage | 1,903 (28.0) | 0.84 (0.77, 0.92) | 0.92 (0.84, 1.01) |
| Other government insurance | 2,220 (30.2) | 0.91 (0.84, 0.99) | 0.92 (0.84, 1.01) |
| Private | 2,349 (33.9) | 1.02 (0.94, 1.11) | 0.95 (0.87, 1.03) |
| Government and private | 1,457 (26.7) | 0.81 (0.74, 0.88) | 0.96 (0.88, 1.05) |
| Uninsured | 796 (49.9) | 1.51 (1.36, 1.67) | 1.22 (1.11, 1.34) |
| Not documented | 147 (45.5) | 1.37 (1.15, 1.63) | 1.08 (0.89, 1.31) |
| Unmet social needs |  |  |  |
| Yes | 811 (35.1) | 1.25 (1.14, 1.36) | 1.13 (1.04, 1.23) |
| No | 4,960 (28.2) | 1 (ref) | 1 (ref) |
| Not assessed | 4,967 (35.2) | 1.25 (1.16, 1.35) | 1.16 (1.07, 1.25) |
| Body mass index |  |  |  |
| <18.5 Kg/m<sup>2</sup> | 120 (34.6) | 1.05 (0.91, 1.23) | 1.04 (0.89, 1.22) |
| 18.5 to <25 Kg/m<sup>2</sup> | 2,461 (32.8) | 1 (ref) | 1 (ref) |
| 25 to < 30 Kg/m<sup>2</sup> | 3,717 (31.4) | 0.96 (0.92, 1.00) | 0.99 (0.95, 1.04) |
| ≥ 30 Kg/m<sup>2</sup> | 4,047 (30.3) | 0.92 (0.89, 0.96) | 0.98 (0.94, 1.03) |
| History of smoking | 4,069 (35.3) | 1.20 (1.15, 1.25) | 1.07 (1.02, 1.12) |
| No history of smoking | 6,602 (29.5) | 1 (ref) | 1 (ref) |
| History of stroke | 1,016 (26.3) | 0.82 (0.76, 0.87) | 0.92 (0.86, 0.98) |
| No history of stroke | 9,722 (32.3) | 1 (ref) | 1 (ref) |
| History of PAD | 771 (22.2) | 0.68 (0.64, 0.73) | 0.87 (0.81, 0.94) |
| No history of PAD | 9,967 (32.6) | 1 (ref) | 1 (ref) |
| History of heart failure | 2,041 (23.6) | 0.69 (0.66, 0.72) | 0.79 (0.75, 0.83) |
| No history of heart failure | 8,697 (34.3) | 1 (ref) | 1 (ref) |
| History of cancer | 954 (25.8) | 0.80 (0.75, 0.85) | 0.91 (0.85, 0.97) |
| No history of cancer | 9,784 (32.3) | 1 (ref) | 1 (ref) |
| Prior CABG | 2,442 (26.4) | 0.79 (0.76, 0.82) | 0.85 (0.81, 0.89) |
| No prior CABG | 8,296 (33.5) | 1 (ref) | 1 (ref) |
| Prior PCI | 7,255 (29.5) | 0.80 (0.77, 0.83) | 0.84 (0.81, 0.88) |
| No prior PCI | 3,483 (37.1) | 1 (ref) | 1 (ref) |
| Diabetes | 4,039 (25.5) | 0.69 (0.67, 0.72) | 0.80 (0.77, 0.83) |
| Without diabetes | 6,699 (36.9) | 1 (ref) | 1 (ref) |
| Colantonio – Cholesterol treatment gaps GWTG |  |  |  |
| Dyslipidemia | 6,836 (26.0) | 0.51 (0.49, 0.54) | 0.61 (0.58, 0.64) |
| Without dyslipidemia | 3,902 (50.8) | 1 (ref) | 1 (ref) |
| Hypertension | 8,577 (29.1) | 0.61 (0.58, 0.63) | 0.86 (0.81, 0.90) |
| Without hypertension | 2,161 (48.0) | 1 (ref) | 1 (ref) |
| **Hospital-level characteristics** |  |  |  |
| **Bed size** |  |  |  |
| Less than 100 beds | 1,261 (43.8) | 1.44 (1.23, 1.67) | 1.16 (0.96, 1.39) |
| 100-199 beds | 2,291 (32.6) | 1.07 (0.95, 1.21) | 1.01 (0.91, 1.13) |
| 200-299 beds | 2,390 (30.5) | 1 (ref) | 1 (ref) |
| 300-399 beds | 1,238 (30.1) | 0.99 (0.84, 1.16) | 0.97 (0.83, 1.14) |
| 400-499 beds | 882 (31.3) | 1.03 (0.88, 1.20) | 0.99 (0.85, 1.15) |
| 500 or more beds | 2,382 (28.4) | 0.93 (0.82, 1.07) | 0.98 (0.85, 1.13) |
| **Accreditation by The Joint Commission** |  |  |  |
| Yes | 8,540 (31.2) | 0.94 (0.85, 1.05) | 1.02 (0.92, 1.14) |
| No | 1,904 (33.2) | 1 (ref) | 1 (ref) |
| **Adult cardiac surgery** |  |  |  |
| Yes | 6,140 (29.8) | 0.84 (0.77, 0.92) | 0.96 (0.86, 1.06) |
| No | 2,875 (35.5) | 1 (ref) | 1 (ref) |
| **Adult cardiology services** |  |  |  |
| Yes | 8,442 (30.9) | 0.73 (0.63, 0.85) | 1.20 (0.93, 1.53) |
| No | 573 (42.1) | 1 (ref) | 1 (ref) |
| **Adult diagnostic catheterization** |  |  |  |
| Yes | 8,158 (30.5) | 0.69 (0.58, 0.81) | 1.15 (0.90, 1.46) |
| No | 857 (44.4) | 1 (ref) | 1 (ref) |
| **Adult interventional cardiac catheterization** |  |  |  |
| Yes | 8,111 (30.4) | 0.67 (0.58, 0.78) | 0.83 (0.68, 1.03) |
| No | 904 (45.1) | 1 (ref) | 1 (ref) |
| **Rural Referral Center** |  |  |  |
| Yes | 3,171 (28.6) | 0.86 (0.79, 0.94) | 0.93 (0.86, 1.01) |
| No | 7,273 (33.1) | 1 (ref) | 1 (ref) |
| **Sole Community Provider** |  |  |  |
| Yes | 333 (33.3) | 1.06 (0.95, 1.17) | 0.82 (0.71, 0.94) |
| No | 10,111 (31.5) | 1 (ref) | 1 (ref) |
| **Teaching Status** |  |  |  |
| Major | 1,422 (27.4) | 0.73 (0.64, 0.83) | 0.91 (0.77, 1.07) |
| Minor | 6,354 (30.6) | 0.81 (0.73, 0.90) | 0.95 (0.85, 1.06) |
| Non-teaching | 2,668 (37.5) | 1 (ref) | 1 (ref) |

CABG: coronary artery bypass grafting; LDL-C: low-density lipoprotein cholesterol; PAD: peripheral artery disease; PCI: percutaneous coronary intervention; ref: reference; SD: standard deviation.
The crude model includes each variable one at a time.
The fully adjusted model includes all variables in the table, simultaneously, and sex, race/ethnicity, and interaction terms between sex and race/ethnicity.

Fully adjusted PRs for an LDL-C ≥70 mg/dL associated with age 70-79 and ≥80 years versus 45-59 years were 0.78 (95% CI 0.75, 0.81) and 0.74 (95% CI 0.71, 0.78), respectively. All other PRs for an LDL-C ≥70 mg/dL ranged between 0.80 and 1.20 (**Supplemental Table 6**).

## Discussion

In the current analysis of the GWTG-CAD registry, almost one-third of patients with CAD hospitalized for a recurrent event were not using a statin. Additionally, approximately three-quarters of patients not using a statin, and half of those using this medication, had an LDL-C ≥70 mg/dL. Statin nonuse was modestly more common among women. Women and non-Hispanic Black and Hispanic patients were more likely to have inadequately controlled LDL-C, particularly among those using statins. The current results indicate a substantial underuse of guideline-recommended statin therapy and a missed opportunity to reduce the burden of CAD events in this high-risk population.

Using data from the National Health and Nutrition Examination Survey (NHANES) 2015-2020, Aggarwal et al. reported that 32.1% of US adults with CAD did not use a statin, and 73.5% had an LDL-C ≥70 mg/dL.^7^ In 2017, 19.9% of ambulatory patients with CAD in the PINNACLE registry were not using a statin.^23^ In 2021, 23.9% of ambulatory patients with cardiovascular disease in the cvMOBIUS2 registry were not using a statin, and 55.5% had an LDL-C ≥70 mg/dL.^24^ The PINNACLE and cvMOBIUS2 registries include patients seen at participating sites with potentially high standards of care, which may explain the lower prevalence of treatment gaps compared to US adults in NHANES. The current analysis included patients hospitalized for a recurrent event, and this may explain the higher prevalence of treatment gaps compared to the PINNACLE and cvMOBIUS2 registries. GWTG-CAD data used for the present analysis were collected approximately 5 years after the publication of the 2018 AHA/ACC/multi-society cholesterol guideline. This suggests that clinical guidelines may be insufficient to break clinical inertia and reduce cholesterol treatment gaps.

Many recurrent CAD hospitalizations in the present analysis could have been prevented with better cholesterol management. Initiation of high-intensity statin therapy in the ∼10,000 patients not using this medication (mean LDL-C ∼100 mg/dL) could reduce LDL-C levels by 50% (∼50 mg/dL or ∼1.28 mmol/L), which could be expected to translate into a ∼30% risk reduction (∼3,000 potentially preventable events).^1,25^ Data on the intensity of statin therapy at admission are unavailable in the GWTG-CAD registry; however, only half of the patients using a statin in the cvMOBIUS2 and GOULD registries were receiving a high-intensity dosage.^24,26^ Compared to moderate-intensity therapy with pravastatin 40 mg/day, high-intensity therapy with atorvastatin 80 mg/day reduces the risk of recurrent myocardial infarction, revascularization, or coronary death by 14% and the risk of unstable angina requiring hospitalization by 29%.^27^ Finally, despite using high-intensity statin therapy, many patients fail to achieve LDL-C levels <70 mg/dL.^24,28^ Adding ezetimibe or a PCSK9i can further reduce the risk for recurrent events in this population.^1,2^ In a prior simulation of patients with a myocardial infarction (22.5% not using a statin, mean LDL-C 92.6 mg/dL), initiation or intensification of statin therapy with the addition of ezetimibe or a PCSK9i when needed to achieve an LDL-C <70 mg/dL and <55 mg/dL was estimated to reduce the risk for recurrent CAD events by 24.3% and 27.6%, respectively.^28^

Over the past 25 years, inpatient quality improvement initiatives, including the GWTG program, have contributed to reducing cholesterol treatment gaps following a CAD hospitalization.^5,8,9^ However, the persistence of cholesterol treatment gaps in ambulatory patients underscores an urgent need for quality improvement interventions in the outpatient setting. Prior studies have shown that LDL-C testing is underused in CAD patients.^26,29^ Increasing LDL-C monitoring may help to break clinical inertia and reduce treatment gaps.^24,30^ Provider and patient education programs, institutional lipid management protocols, and follow-up by a cardiologist are other interventions that can reduce cholesterol treatment gaps in the ambulatory setting.^26,30–32^ Initiated in 2021, the AHA National Integrated Atherosclerotic Cardiovascular Disease (ASCVD) Initiative is a quality improvement program designed to optimize LDL-C control in ambulatory ASCVD patients through better inpatient-outpatient coordination for lipid monitoring and management.^33^ In patients participating in this initiative, LDL-C ≥70 mg/dL decreased from 51.6% in 2022 to 50.5% in 2023 (p-value <0.005).

The current results suggest the persistence of sex and racial/ethnic disparities in cholesterol management.^10–13,24,26,30^ Statin nonuse and LDL-C ≥70 mg/dL were more common in women compared to men. LDL-C ≥70 mg/dL was also more prevalent in non-Hispanic Black and Hispanic patients using a statin compared to their non-Hispanic white counterparts. However, statin nonuse was lower among non-Hispanic Black versus non-Hispanic white patients. Prior analyses of ASCVD patients in the PALM registry and in the Medicare and MarketScan databases found no association between race/ethnicity and receipt of recommended statin therapy intensity after adjustment.^6,31^ A potential explanation for racial differences in statin nonuse observed in the current analysis is the selection of individuals with a recurrent CAD hospitalization rather than the entire CAD population. In the REasons for Geographic And Racial Differences in Stroke (REGARDS) study, Black participants who discontinued statin were more willing to reinitiate this therapy compared to their white counterparts (age-sex-adjusted PR 1.29, 95% CI 1.05, 1.59).^34^ In addition, Black men were less likely to experience non-fatal myocardial infarction and more likely to have fatal coronary events than white men, potentially due to the underdetection or delayed recognition of symptoms among the former.^35^ By selecting individuals with a recurrent CAD hospitalization rather than all CAD patients, the current analysis may include an overrepresentation of non-Hispanic Black patients with more proactive health-seeking behaviors, including both early recognition of coronary symptoms and high willingness to use statins (i.e., collider stratification bias).^36^

Most patient characteristics in the current exploratory analyses showed a small association with cholesterol treatment gaps, indicating that treatment gaps are not limited to specific subpopulations. After multivariable adjustment, being uninsured was associated with a 22% higher likelihood of statin nonuse. Lowering economic barriers may improve statin adherence and reduce the risk for recurrent events.^37,38^ History of diabetes, dyslipidemia, and heart failure was associated with lower statin nonuse, potentially due to a higher perceived need for statins in these populations. Prior studies in the cvMOBIUS2, GOULD, and PALM registries have consistently shown better cholesterol management in patients with diabetes.^24,26,31^ Reasons for better cholesterol management in people with diabetes may be multifactorial, including patient-related (e.g., treatment adherence) and provider-related factors (e.g., more frequent visits and cholesterol monitoring; better communication).^39,40^ An LDL-C ≥70 mg/dL was less common in older adults, which may reflect a higher frequency of catabolic states, frailty, or malnutrition.^41^

The current analysis used standardized data from a large sample of patients and hospitals from all US census regions. However, this study has limitations. The GWTG program is a quality improvement initiative. Therefore, results may not be representative of all US patients. Data on statin intolerance were unavailable. However, only 6.4% of patients who were discharged alive to their homes were not prescribed this medication. The current analysis included patients hospitalized for a recurrent event; therefore, collider stratification bias cannot be excluded. Some questions in the GWTG registry are optional, and reasons for noncompletion may be unrelated to participant characteristics (i.e., not at random).

Among patients with known CAD hospitalized with ACS, ambulatory cholesterol management remains suboptimal. Disparities in LDL–C levels by sex and race/ethnicity persist, especially among patients using statins. Targeted quality improvement initiatives that promote LDL–C monitoring and timely therapy intensification may help reduce these gaps and improve secondary prevention outcomes.

## Data Availability

Data from the Get With The Guidelines - Coronary Artery Disease registry used in the current analysis were provided by the American Heart Association through the Precision Medicine Platform (https://pmp.heart.org/). These data are available upon appropriate request to the American Heart Association Precision Medicine Platform. The Precision Medicine Platform is powered by Amazon Web Services and supported by Hitachi Vantara. The aims, methods, analytical plan, code for derivation of study variables, and the final sample size for the current analysis are available on the Open Science Framework website (https://osf.io/nd2u9/overview), which is hosted by the Center for Open Science. Other study information is available from the corresponding author.

https://pmp.heart.org/

## Acknowledgements

Data from the Get With The Guidelines – Coronary Artery Disease registry was provided by the American Heart Association.

